# Service evaluation of “GP at Door” of Accident and Emergency Services in Eastern England

**DOI:** 10.1101/2023.09.09.23295296

**Authors:** Julii Brainard, Aiden Rice, Gareth Hughes, Paul Everden

**Affiliations:** Norwich Medical School, University of East Anglia Norwich NR4 7TJ, UK; North Norfolk Primary Care, Alkmaar House, Alkmaar Way, Norwich NR6 6BF, UK

**Author notes:** Corresponding author. Postal Address as above.

**Keywords:** <u>Keywords</u>: Accident and Emergency, low acuity presentations, wait times, service evaluation

## Abstract

**Aims:** To describe activity and outcomes after streaming low urgency attenders to general practice services at door of Accident and Emergency departments (GDAE), including possible benefits to co-located emergency departments.

**Methods:** As a service evaluation, we describe GDAE users, their reasons for presentation, wait times, outcomes and co-located emergency department performance metrics at two hospitals in eastern England.

**Findings:** Each GDAE saw about 928 patients per month. Wait times for usual A&E care relatively shortened at only one site. Reattendances were common (about 10% of attenders), 75% of GDAE attenders were seen within 1 hour of arrival, 7% of patients initially allocated to GDAE were referred back to A&E for further investigations, 59% of GDAE patients were treated and discharged with no further treatment or referral required. Pain, injury, infection or feeling generally unwell each comprised > 10% of primary reasons for attendance. Referrals to specialist health services were outcome for 4% and 16% at respective sites.

**Conclusions:** About 26,000 A&E attendances appear to have been prevented. Patients were seen quickly at both GDAE sites, while there were more specialist referrals or shorter wait times for usual A&E services at only one site. Process evaluation could illuminate reasons for these differences.

## Introduction

Attendances to UK National Health Service (NHS) accident and emergency departments (A&Es) have risen steadily since the 1990s, for diverse and many reasons. [1] In 2004, NHS A&Es were set a target (key performance indicator, KPI) to admit, transfer for treatment or discharge at least 95% of attending patients within four hours of presentation (4hKPI). [2] Waits above 5 hours in A&E have been linked to higher mortality within 30 days after attendance [3] as well as longer hospital stays for patients admitted from A&E. [4] Most NHS A&Es have failed the 4hKPI target in most time periods, since 2016. [5] There is thus huge ongoing interest in innovative ways to better manage the needs of persons attending A&E to try to help A&Es achieve fast care delivery and reduce risk of health harms associated with delays to treatment or inappropriate treatment.

Previous observations [6–9] suggested that 15-44% of Emergency Department attenders in Britain sought health care for conditions that are suitable for other health services, especially primary care, where care could be administered by general practitioners (GPs) themselves and/or advanced nurse practitioners (ANPs). Putting A&E attenders in need of GP-level treatment on a more appropriate pathway seems likely to produce many benefits: fewer breaches of the 4hKPI, faster resolution of health complaints for all, more proportionate investigation and treatment pathway, and better chances of continuity of care. [8] During the COVID-19 pandemic, shorter waiting times and faster treatment was also desirable to reduce risk of nosocomial COVID-19 transmission. [10]

Since 2011, it is increasingly common for GPs to be incorporated into A&E streaming and for GP-services to be sited as distinctive service areas within or adjacent to NHS A&Es. [11] NHS five year planning [12] stated that every A&E should put in place ‘front-door clinical streaming’, to quickly find the most appropriate pathways for A&E attenders. One way to implement such streaming is via an immediately available GP-level service as alternative to A&E. Here we evaluate programmes that offered the full range of usual GP services to walk-in patients at two acute care providers in England. In the context of a service evaluation study design, using routinely collected and published data, we consider aspects of service provision including: activity (counts of presentations), socio-demographic profiles of service users, reasons for presentation, movement to a more urgent pathway, service user satisfaction, data about reattendances and concurrent performance of the 4hKPI in co-located A&E compared to similar A&Es elsewhere in England.

## Methods

### Service Description

GDAE services were commissioned by the Norfolk and Waveney integrated care system (N&WICS), in coastal eastern England. 97% of hospital attendances by the approximately one million patients registered with N&WICS are at three acute care providers. [13] A pilot GP at Door of A&E (GDAE) service ran at the region’s largest acute care centre (Norwich and Norfolk University Hospital, NNUH) from December 2019 to February 2020 and is described elsewhere. [14] The GDAE service hoped to provide patient and system benefits: converting unplanned to planned treatment; providing clinician access to full primary care record which was used in assessment; primary care records to be updated immediately; reduction in unnecessary investigations and more appropriate risk management.

GDAE services were initiated and replicated in the NNUH format at the other two secondary care providers in N&WICS in late 2021/early 2022: Queen Elizabeth Hospital (QEH) and James Paget University Hospital (JPUH). The GDAEs at JPUH and QEH ran 7 days/week, 9am- 9pm. Clinical GDAE staff were typically at one GP and one ANP. Data about the GDAEs were available from service initiation, 16 months at the JPUH (October 2021-January 2023) and 12 months at the QEH (Feb 2022-January 2023).

The GDAE care pathway is illustrated and described at length elsewhere. [14] Patients attending the A&E walk-in entrance (arrival not by ambulance) were met by a GP or ANP who triaged them to the GDAE service or usual A&E care. Bespoke triage criteria were applied according to criteria set out in the Appendix and relying on clinical acumen. Patient care during this attendance was meant to be identical to care available at their own registered GP surgery. GDAE patients were booked in and asked for consent to access their primary care records. Care was provided by a GP or ANP and primary care records updated accordingly. Transfer back to the usual A&E pathway was possible at any time.

### Activity and Outcomes

These services were described and evaluated using the data and outcomes listed below, from data routinely collected.

-- Service user descriptions (sex, age, deprivation levels)

-- Total GDAE activity (counts of attendances, appointments, and dispositions)

-- Wait times

-- Outcome after completing GDAE appointment

-- Frequency of reattendance

-- Most common reasons for attendance to GDAE, including among persons who reattended during the monitoring period

-- How well each of the co-located A&Es concurrently met 4hKPI compared to historical data or similar A&Es

### Data: Service Users

Data were routinely collected in electronic medical records (SystmOne and Symphony). The data described individual GDAE attenders resident in both N&W and out of area, using the below fields:

-- Unique patient identifier

-- GDAE Location

-- Date of attendance

-- Time at booking in

-- Appointment status (eg., finished or cancelled)

-- Gender

-- Age

-- IMD2019 group quintile

-- Reason for attending

-- Treatment outcome (eg., referral, discharge or not recorded)

-- Wait from arrival time to time seen (minutes)

The Index of Multiple Deprivation 2019 (IMD2019) [15] is a national ranking of relative deprivation in residential areas. These ranks were available in five ordinal categories (1 = most deprived to 5 = least deprived) relative to all-England. Available (in system coding at time of booking) reasons for attendance (descriptions of primary complaint when patient presented) are listed in Box 1.

**Box 1.**
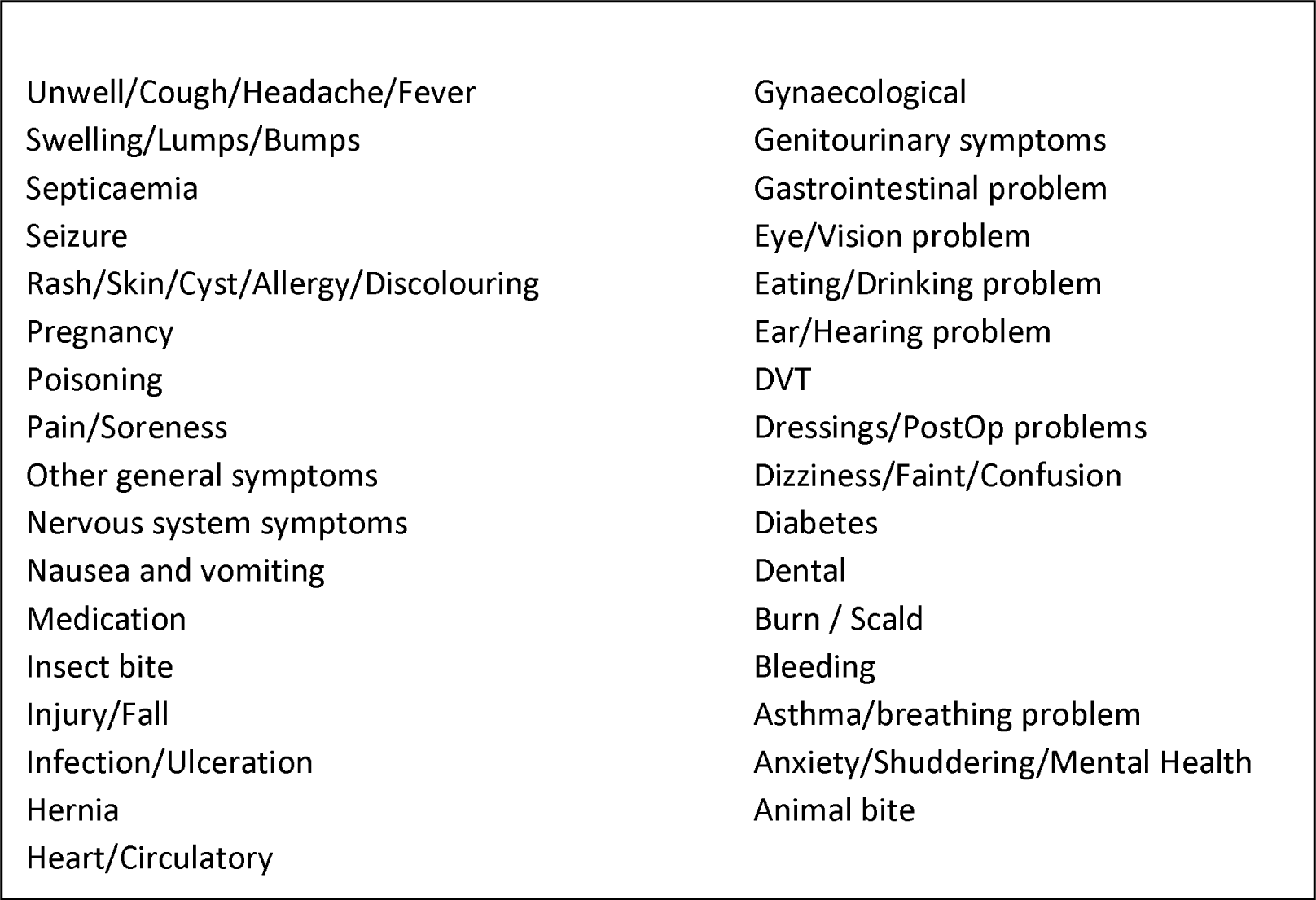
Primary reasons for attending A&E, as recorded at GDAE booking N&W A&Es.

### Data: Four hour target for discharge or admission for NHS A&Es

NHS England publishes statistics (percentages) for how many patients completed their A&E visit within four hours. Just prior to GDAE initiation, in September 2021, the JPUH A&E had 7135 completed attendances, the QEH had 6487 attendances. From NHS England datasets, we extracted 4hKPI values for prior 12 and subsequent months after GDAE services started at JPUH and QEH, as well as for all (n=23) comparator type 01 (full range of urgent care services provided) NHS emergency departments that had somewhat similar attendances (between 4000 and 10,000) in September 2021. The 23 comparator sites are listed in the Appendix. Many initiatives to try to improve the 4hKPI are happening widely in the NHS, although we do not know their full extent at each of these specific 23 comparator sites. Rather, our rationale was to try to explore whether the 4hKPI at JPUH and QEH had improved, deteriorated or remained static compared to KPI performance over a similar period at comparable NHS A&Es. We also compared the 4hKPI at JPUH/QEH, pre- and post-implementation of their GDAE services. We looked at overall 4hKPI at both sites and disaggregated wait times by major and minor acuity problems at JPUH; this disaggregation was unavailable for the QEH.

### Service user satisfaction

We summarise responses to an online survey that service users were invited to take; survey questions are in the Appendix.

### Analysis

Descriptive statistical analysis, including comparisons between the JPUH and QEKL using tests of proportionality or distribution difference: chi square, Kolmogorov-Smirnov and Mann Whitney U test. Significance threshold was p < 0.05. Concurrent statistics for the 4hKPI are provided and discussed narratively before and after the N&W GDAE services were deployed (first 12 months only), comparing JPUH and QEH with their own historical data and similar A&E departments in England.

## Results

### Service overview: Activity, users and their outcomes

Table 1 shows overview information for each and both services. Over 99% of patients booked into GDAE attended their appointments. Other bookings did not proceed due to cancellation (by user or service) or when patient did not attend. Detailed demographics and outcomes are provided for only patients who completed appointments; denominators for percentage calculations exclude records where an attribute was not recorded. Median age and age distribution of attenders were nearly identical at the sites (median = 33 years, range 0-100). Items in bold font in Table 1 had significant proportional differences (chi square test) between JPUH and QEH. JPUH patients were significantly more likely to come from the most deprived quintile areas, 36.9% vs. 26.1% at QEH. QEH had more attendances by patients resident outside Norfolk and Waveney. 12.6% of JPUH service users made repeat attendances, compared to 9.3% of QEH patients. The most GDAE attendances by an individual at each site was 14. The proportions of several treatment outcomes differed significantly between sites in some respects: QEH patients were more likely to be referred to other services (A&E or specialisms) or to be referred back to their GP with recommendations for further treatment/investigations.

**Table 1.** Service activity and users overview.

|  | JPUH: n, % | QEH: n, % | All: n, % |
| --- | --- | --- | --- |
| Appointments made | 18,212 | 10,656 | 28,868 |
| Appointments attended | 18,087, 99.3% | 10,627, 99.7% | 28,714, 99.5% |
| *Of those who completed an appointment with GDAE ...* |  |  |  |
| Females | 9276, 51.2% | 5578, 52.5% | 14,854, 51.7% |
| Median Age (IQR) | 33, 14-55 | 32, 12-55 | 33, 13-55 |
| In most deprived quintile | **6658, 36.9%** | **2752, 26.1%** | 9410, 32.8% |
| Resident in N&W | **16,763, 92.2%** | **7683, 72.3%** | 24,446, 85.1% |
| *Reattendance* |  |  |  |
| Unique patients | 15,398 | 9534 | 24,932 |
| # Patients who made 2 visits | **1484, 9.6%** | **726, 7.6%** | 2210, 8.9% |
| ... 3 visits | **311, 2.0%** | **93, 0.98%** | 404, 1.6% |
| .... 4 visits or more | **149, 0.97%** | **48, 0.50%** | 197, 0.79% |
| *Outcome after being seen* |  |  |  |
| *Discharged after...* |  |  |  |
| Treatment with no further treatment required | **11,629, 64.4%** | **5220, 49.2%** | 16,849, 58.8% |
| Not treated with no further treatment required | 2618, 14.5% | 1468, 13.8% | 4086, 14.3% |
| Unclear if treated, but no referral or further treatment required | 583, 3.2% | 303, 2.9% | 888, 3.1% |
| *Referred to...* |  |  |  |
| GP, further action recommended | **1423, 7.9%** | **904, 8.5%** | 2327, 8.1% |
| GP, for watchful waiting | 21, 0.1% | 1, 0.01% | 22, 0.1% |
| Specialty service | **770, 4.3%** | **1706, 16.1%** | 2476, 8.6% |
| A&E immediately | **1007, 5.6%** | **1007, 9.5%** | 2014, 7.0% |
| Patient declined further treatment or referral | 3, 0.02% | (none) | 3, 0.01% |

Note: Outcomes were mutually exclusive. If patients were treated prior to a referral was not recorded. Items in bold were significantly different between JPUH and QEH at $p \leq 0.05$ .

**Table 2.** Primary reason for attendance to GDAE services.

| Reason for attendance | All attendances |  | Reattenders |  |
| --- | --- | --- | --- | --- |
|  | JPUH<br>n=17295 | QEH<br>n=10606 | JPUH<br>n=1945 | QEH<br>n=867 |
| Animal bite | 2.0% | < 1 % | 1.5% | < 1 % |
| Asthma/Breathing | 2.6% | 2.1% | 3.1% | 2.3% |
| Dental | 6.1% | 4.6% | 6.1% | 4.4% |
| Dressings/PostOp problems | 2.7% | 1.2% | 3.9% | 2.3% |
| DVT | < 1% | 1.5% | < 1 % | 1.0% |
| Ear/hearing problem | 3.4% | 3.7% | 4.8% | 5.0% |
| Eye/Vision problem | 4.0% | 7.3% | 3.1% | 5.1% |
| Gastrointestinal problems | 4.0% | 5.5% | 3.8% | 6.3% |
| Gynaecological | 1.1% | 1.7% | 1.3% | 1.6% |
| Infection/Ulceration | 11.4% | 9.6% | 13.3% | 11.4% |
| Injury/Fall | 13.3% | 12.3% | 8.2% | 7.5% |
| Insect bite | 1.3% | < 1% | 1.1% | < 1 % |
| Nervous system symptoms | < 1% | 1.2% | < 1 % | 1.2% |
| Pain/Soreness | 17.9% | 16.9% | 15.7% | 14.0% |
| Rash/Skin/Cyst/Allergy/Discolouring | 4.5% | 4.1% | 5.4% | 6.5% |
| Swelling/Lumps/Bumps | 1.2% | 1.7% | 1.2% | 1.7% |
| Unwell/Cough/Headache/Fever | 10.2% | 13.5% | 12.8% | 18.3% |
| Other stated conditions (each <1%) | 14.4% | 13.1% | 14.6% | 11.3% |

Note: Totals are for visits with presenting complaint recorded; there were additional visits without reason for presentation recorded. PostOp = postoperative problems.

### Timing of attendance and waits to be seen

For patients who completed appointments, Figures 1a-1c depict these aspects of service attendance: arrival time (resolved to nearest hour); month of attendance (last/concurrent 12 months only); wait from arrival time to time seen (whole minutes). There was negligible difference between sites with respect to proportion of attendances in any specific calendar month (Figure 1b). However, both times of arrival (1a) and wait times (1c, from arrival to being seen) were different between sites. 55.6% of QEH patients arrived by 2pm, compared to 48.9% of JPUH patients (difference significant at p < 0.001, Kolmogorov-Smirnov test). Elapsed time from arrival to being seen was greater at QEH (p < 0.001, Mann Witney U test), with QEH median elapsed time = 26 minutes (IQR 8-51), while the time from arrival to being seen at JPUH was median 22 minutes (IQR 7-45). Durations of actual appointments at both sites were extremely similar: QEH median 15 (IQR 11-21) and JPUH median 16 (IQR 11-22). We also looked at proportions arriving by day of week (eg., Monday, Tuesday…) and found no significant difference between sites (data not shown).

**Figure 1.**
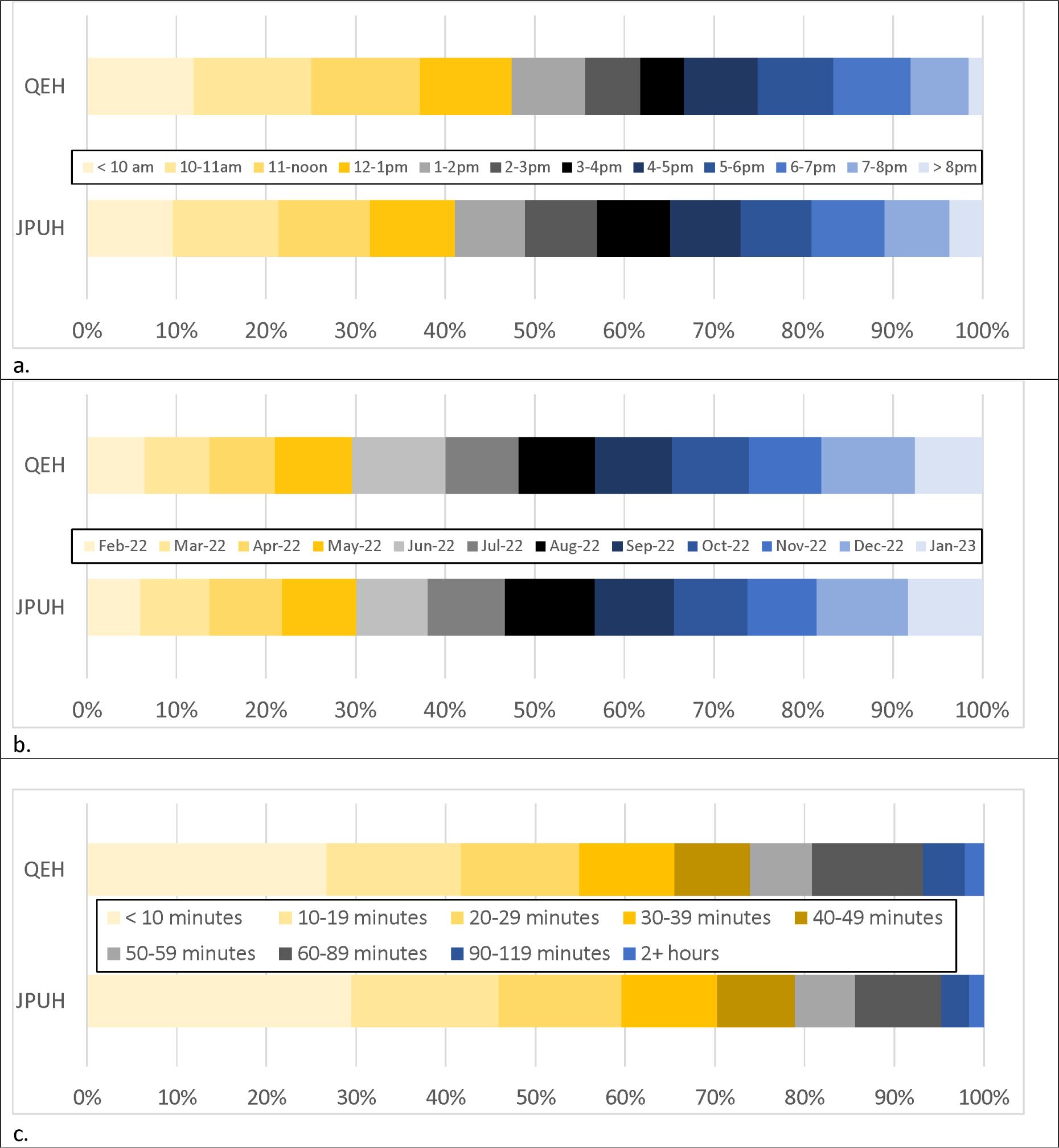
Service activity for completed GDAE service users. a. time of day for arrivals; b. arrival proportions in most recent 12 months of service; c. elapsed time from arrival to being seen.

### Reattendance and reasons for presentation

Primary reasons for presentations that comprised at least 1% of visits are listed individually in Table 2; all other reasons (such as ‘diabetes’) each comprised < 1% of visit reasons each and are grouped under “Other conditions”. 925 persons attended the QEH GDAE more than once in the 12 months of monitoring, 1945 persons reattended the JPUH GDAE more than once in 16 months of service operation. Persons who attended more than once had a very similar distribution of reasons as single occasion attenders. The largest difference between one-time and repeat attenders, is that reattendances at QEH were much more likely to be for feeling ‘generally unwell’ : 18.3% among repeat attenders vs. 13.5% for full group.

**Table 2.** Results of patient satisfaction survey.

| Question | JPUH | QEH | Combined |
| --- | --- | --- | --- |
| *Who referred you to A&E today?* |  |  |  |
| No one/ brought self | 61% | 73% | 65% |
| NHS111 (telephone/online advice service) | 22% | 6.7% | 15% |
| GP advised attendance | 9.8% | 20% | 14% |
| Chemist/pharmacist | 2.4% | 0% | 1.4% |
| Nurse in A&E | 2.4% | 0% | 1.4% |
| St. John Ambulance | 2.4% | 0% | 1.4% |
| *Would you recommend the service to friends and family?* |  |  |  |
| Yes | 90% | 97% | 93% |
| No | 10% | 3% | 6.9% |
| *Were you aware of this service prior to arrival?* |  |  |  |
| Yes | 15% | 23% | 18% |
| No | 85% | 77% | 82% |
| *How did you find person who met you at the entrance ...* |  |  |  |
| Good or Excellent | 86% | 96% | 90% |
| Fair | 5% | 3% | 4% |
| Poor or Very Poor | 2% | 0% | 1% |
| *Booking receptionist was ...* |  |  |  |
| Very helpful | 93% | 97% | 94% |
| Somewhat helpful | 5% | 0% | 2.8% |
| Neither helpful nor unhelpful | 2% | 0% | 1.4% |
| Unhelpful or very unhelpful | 0% | 0% | 0% |
| No receptionist only clinicians at arrival | 0% | 3% | 1.4% |
| *How did you find consultation with clinician ...* |  |  |  |
| Good or Excellent | 95% | 100% | 97.2% |
| Fair or Poor (user entered both answers) | 2.4% | 0% | 1.4% |
| Poor | 2.4% | 0% | 1.4% |
| Very Poor | 0% | 0% | 0% |
| *How do you think we could improve the services?* |  |  |  |
| No comment/Nothing needs improvement | 76% | 67% | 72% |
| Wait times could improve | 7.1% | 6.7% | 6.9% |
| Should be publicised more | 2.4% | 13% | 6.9% |
| Other specific comments or suggestions | 14% | 13% | 14% |

### 4 hour target statistics, study sites and in similar A&Es

Figure 2a illustrates overall 4hKPI performance at JPUH and QEH A&Es each month, compared to a group of similar size (activity level) A&Es in England that also provide a full range (Type 1) of emergency department services. On Figures 2a and 2b, the 4hKPI of median performing comparator A&E is indicated with the central black line. The full range of comparator 4hKPIs is denoted by grey shaded areas. Further disaggregation was available at JPUH to generate Figure 2b (majors treatment streams at JPUH and comparator A&Es), while comparisons for the minors patients is described below in text. Information for June 2021 is interpolated on the majors chart because only two relevant A&Es reported majors 4hKPIs this month. On Figures 2a and 2b, dashed lines indicate the period prior to the introduction of the GDAE services at JPUH / QEH (minimum 12 months before). The GDAE services did not coincide with consistently improved overall 4hKPI at JPUH or QEH compared to their own preceding 12 months of data. However, it makes more sense to consider the 4hKPI relative to the comparator group of A&Es, given that there is long term deterioration in 4hKPI at all NHS A&Es, a decline which has accelerated since 2019. [16] For attendances with minor acuity (“minors”), JPUH 4hKPI statistics prior to GDAE service were median 93.9% (range 48.6-100%). After GDAE started the JPUH minors 4hKPI was median 99.85% (range 99.3-100%). For comparator A&Es throughout the 28 month period, the minors 4hKPI among comparator A&Es was median 99.9% (range 61.3-100%). The JPUH 4hKPI for minors was much closer to the comparator performance in the period after GDAE was introduced. This result is considered with caution because while missing data were unusual for majors or overall 4hKPI, the minors-only 4hKPI was often available for only 10-15 of 23 possible comparator A&Es.

**Figure 2:**
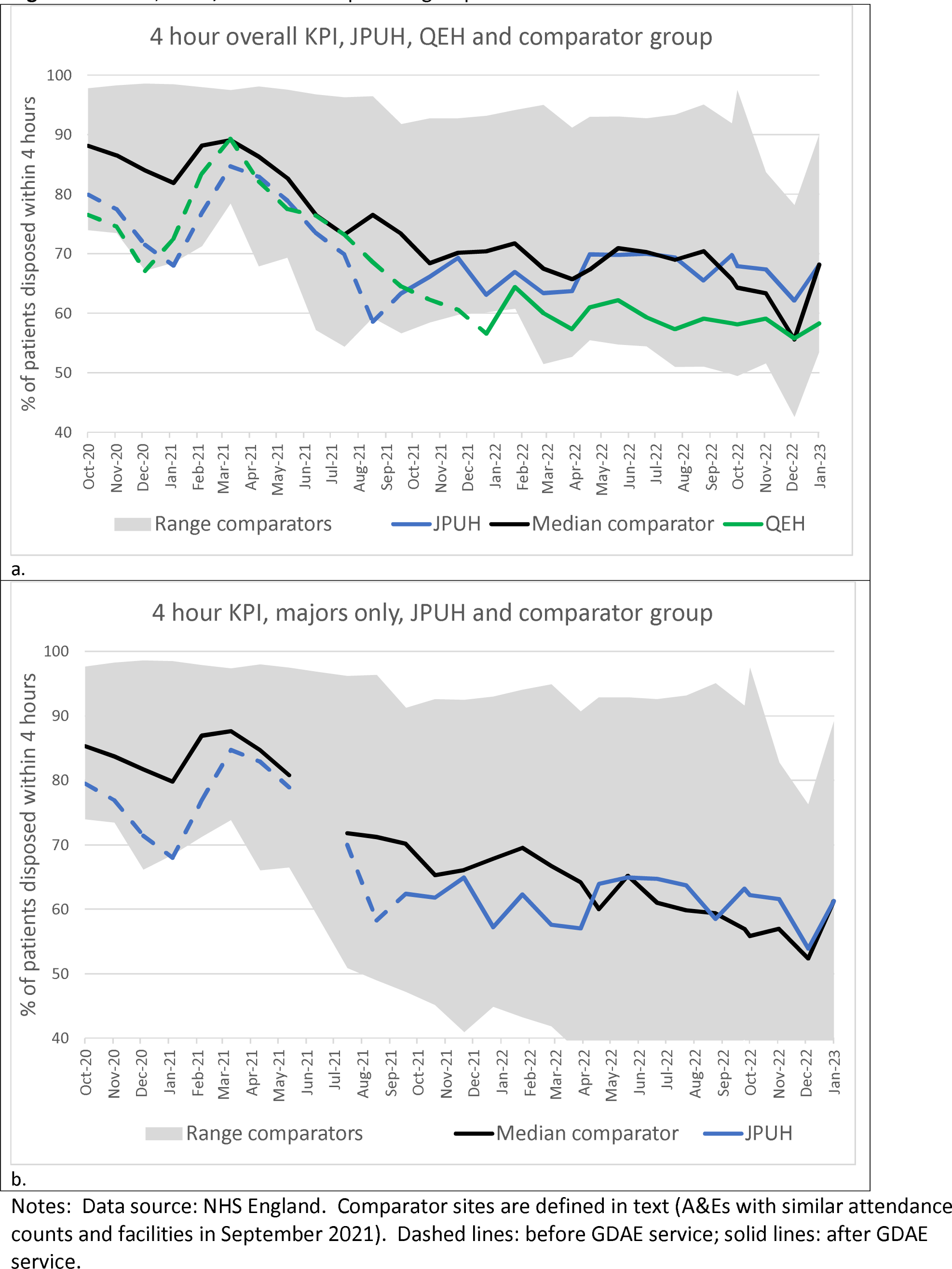
Four hour key performance indicator (4hKPI) at JPUH/QEH and comparator group of Accident and Emergency departments

Compared to similar A&Es, the JPUH and QEH were consistently in the 50% of lower performing A&Es prior to GDAE introduction. After GDAE, the JPUH service had consistent improvement relative to the comparators in 4hKPIs, overall (Figure 2a), for majors (Figure 2b) and for minors (data stated in text). In contrast, the QEH was among the 25% of lowest performing A&Es in this group of A&Es before and after GDAE. These data do not show that, compared to a diversity of other wait-reduction initiatives that other similar A&Es must have implemented, that GDAEs have delivered consistent 4hKPI improvements.

### Service user satisfaction

42 patients at JPUH and 32 patients at QEH gave data about patient satisfaction (Table 2). There were many very positive comments about care received, naming individual clinicians and specific points of gratitude. Most respondents said that there was nothing they could think of to improve the service. There were three negative statements in the open text responses: 1) at-door streamer made someone feel like a time-waster; 2) excess walking distance from A&E entrance to entrance to GDAE; 3) brusque manner of consultation GP; all negative comments were at the JPUH.

## Discussion

There is a long-standing objective (NHS [12]) that every A&E should put in place ‘front-door clinical streaming’, to quickly find the most appropriate pathways for A&E attenders. One way to implement such streaming is with use of an immediately available GP-level service.

Over 26,000 attendances at these study sites were diverted to GDAE away from A&E. Recent other analyses concluded that primary care clinicians working in A&Es did not increase efficiency of services, improve clinical outcomes, patient or staff experiences. [17, 18] However, those studies did note that there is a large variety in how primary care clinicians work at A&Es [19], with likely resulting variations in benefits that may result. Our findings for “full-service” GP services co-located at two A&Es were that the 4hKPI improved in several domains concurrent with GDAE service deployment at the JPUH, but not at the QEH. The reasons for this difference probably requires process evaluation [20] to thoroughly understand, and to inform whether the GDAE service should be credited or if other factors led to strong relative improvement at JPUH but not QEH. For instance, management of admission bed availability that differed between the sites may have had the greatest impact on 4hKPIs. Concurrently during this period the QEH had to contend with failing infrastructure which may have undermined efficacy and quality of services. [21] Also relevant may be somewhat different aspects between the sites with respect to times of day when patients attend or their demographic profiles. We intend to explore GDAE attender demographic traits in a separate analysis.

We did not observe strongly different reasons for attendance among service users between sites or between single-occasion attenders versus reattenders. Clinical audits and process evaluation would also be helpful to understanding why QEH patients were much more likely to be referred to other services.

We plan to undertake future research to better understand barriers faced by N&W residents when trying to obtain GP-level care. We have used involvement of patient and public advisors to explore how A&E attenders with low acuity conditions may be asked about their experiences of seeking health care without conferring stigma. Collecting such data would provide specificity and focus about real-world barriers that people face and cause them to find it appealing to obtain unscheduled GP-level care by attending A&E. Preliminary comments from our public advisors include that from a patient’s perspective, A&E visits seem to provide more reassurance and faster resolution than visits to a GP surgery. GDAE format services may help to provide these benefits to persons who attend A&Es while reducing the risks of possible over-investigation and over-treatment [22], especially in an era when routine GP appointments seem very challenging to obtain. [23]

## Strengths and Limitations

To know whether the GDAE services described in this service evaluation have achieved the most appropriate level of care would require patient history audits by clinically qualified staff; we did not have resources to attempt that. We lacked access to sensitive individual patient clinical data to assess how often reattendances were for the same condition that caused initial presentation. We recommend to commissioners that evaluation of potential benefits or harms of GDAE format services should ideally include clinical audit to ascertain if reattendances tend to be for exacerbations of initial conditions or for unrelated health problems. Such audit results would, however, need to be interpreted with reference to frequency of reattendance to A&E itself for related conditions, and/or to community GP surgeries, for escalations of health problems that were recently treated by same type of service.

Performance of the 4hKPI at JPUH and QEH was only analysed descriptively; a different study design would be necessary to evaluate GDAE design services impacts on the 4hKPI robustly against other initiatives that try to improve the 4hKPI. We have not attempted cost benefit analysis; it seems likely that most GDAE services do not provide high value for money because of their frequent use of locum staff (paid at higher rates than contracted staff). Our study benefited from a large and comprehensive dataset because of careful planning from time of service inception to ensure an informative service evaluation. “Lack of consistent evaluation” is a chronic problem in the NHS [24], which means that commissioners struggle to know what benefits may have been achieved or not; our study attempts redress this information deficit.

GP at door of A&E services in N&WICS appeared to avert more than 26,000 A&E attendances. This may have helped cause a relative improvement in the 4hKPI at one of the study sites. The services have been busy and resulted in high satisfaction among service users. The JPUH service was somewhat more efficient than QEH (shorter wait times, fewer onward referrals) in spite of JPUH serving a more deprived (presumably with greater morbidities) population; this finding suggests there may be opportunities for efficiency gains at the QEH GDAE. Reattendance is common at both sites, suggesting an undesirable outcome : some attenders may now prefer GDAE over their usual GP service. GDAE data could be used to help frequent attenders find more appropriate service pathways. Process evaluation would be useful to understand why GDAE seemed more successful at one site than the other.

## Data Availability

Some of the data are available publicly from NHS, secondary analysis. Other (patient and service) data are not available publicly.

## Acknowledgements

The services were hosted by the JPUH and QEHKL and funded by Norfolk & Waveney integrated care system and JPUH. JB is affiliated to the National Institute for Health Research Health Protection Research Unit (NIHR HPRU) in Emergency Preparedness and Response at King’s College London in partnership with the UK Health Security Agency (UK HSA) and collaboration with the University of East Anglia. The views expressed are those of the authors and not necessarily those of any collaborating or funder institution.

## Conflict of interest

All authors declare that we have no conflict of interest. Service activity data were collated by a contracted third party organization, Paradigm Leaders, who had no role in data analysis or writing this document or its conclusions.

Ethical approval for this study was granted by the University of East Anglia Faculty of Medicine and Health Ethics Committee (Reference: ETH2122-1954, 22 June 2022).

## Author contributions

PE helped design the service and conceived of the evaluation, which was designed by JB and PE. AR and JB designed data collection and secured ethics and research governance approval with oversight from GH. JB and AR planned and undertook analysis, JB wrote the first draft and assembled revisions. All authors revised for content.

## Funding

The services were hosted by the JPUH and QEHKL and funded by Norfolk & Waveney integrated care system and JPUH. Dr. Brainard is affiliated to the National Institute for Health Research Health Protection Research Unit (NIHR HPRU) in Emergency Preparedness and Response at King’s College London in partnership with the UK Health Security Agency (UK HSA) and collaboration with the University of East Anglia. The views expressed are those of the authors and not necessarily those of any collaborating or funder institution.

Author contributions:

PE helped design the service and conceived of the evaluation, which was designed by JB and PE. AR and JB designed data collection and secured ethics and research governance approval. JB and AR planned and undertook analysis, JB wrote the first draft and assembled revisions. All authors revised for content.

Service evaluation of “GP at Door” of Accident and Emergency Services in Eastern England

## Appendix

### Triage Inclusion/Exclusion criteria

The service does not have a strict inclusion/exclusion criterion. It relies on the clinical acumen of the clinicians working in the service. The criteria that are in place are outlined below:

### Patient Inclusion Criteria

* All patients who self-present to the emergency department will have the opportunity to access a general practice appointment if they are registered with a Norwich, North Norfolk, West Norfolk, Great Yarmouth and Waveney, or South Norfolk GP Practice and are assessed by a clinician to be suitable for primary care.
* Patients out with this catchment area may also be seen by the service due to an update in the process and agreement. Details of their review by the service will be sent to their own GP surgery.
* Children may be seen by the service for minor illness complaints where appropriate as assessed by the GP or ANP.

### Patient Exclusion Criteria

* Any patient requiring intervention or investigation within ED
* Non-traumatic chest pain
* Any patients with signs or symptoms of stroke
* Any patients, that are not registered with a GP Surgery within the UK
* All children presenting to ED with injuries
* Children under three months

Study sites (yellow fill) and comparator sites (green fill) in England, UK. Comparator sites were chosen because they had between 4000 and 10,000 Type 01 attendances in September 2021.

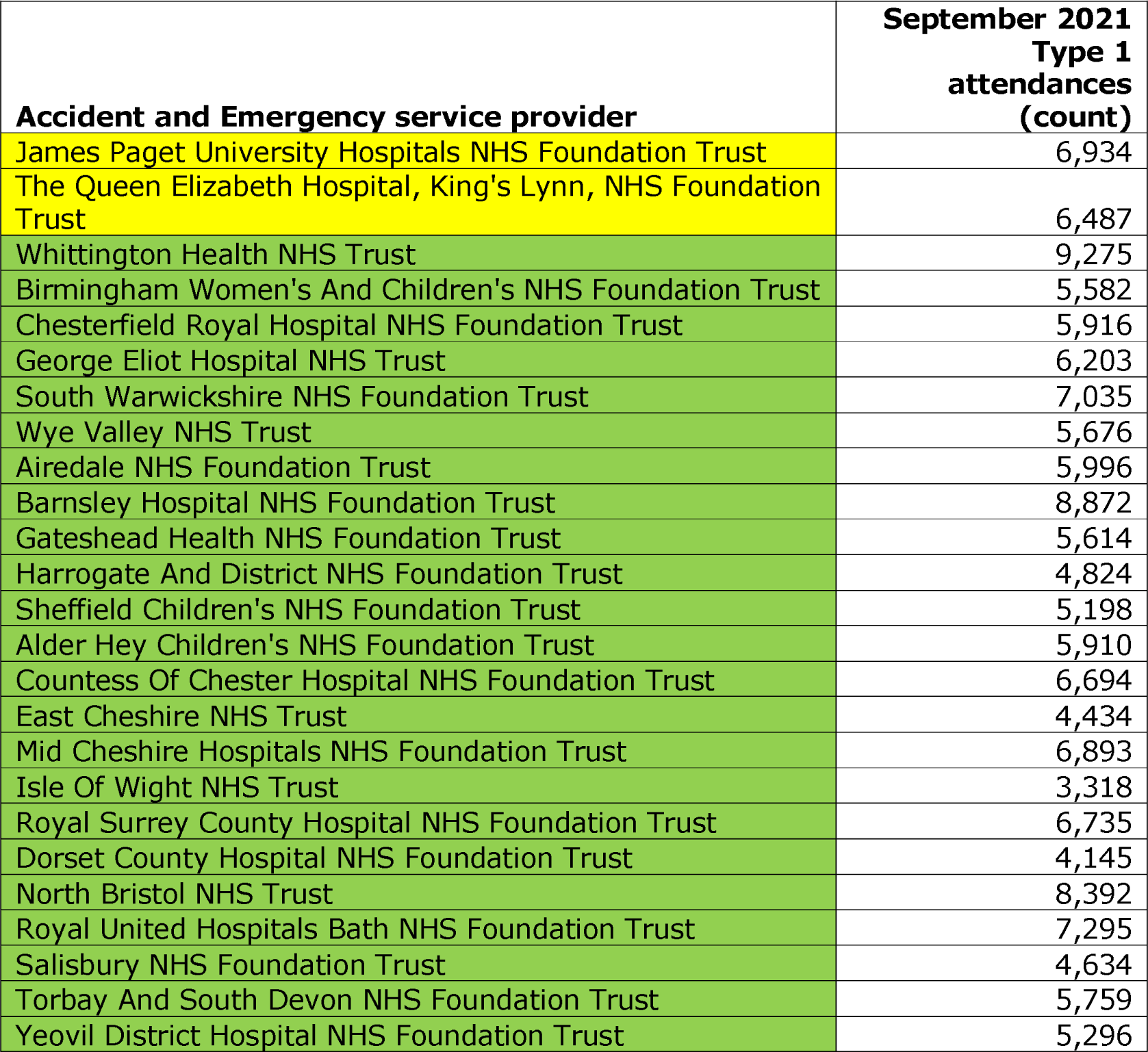
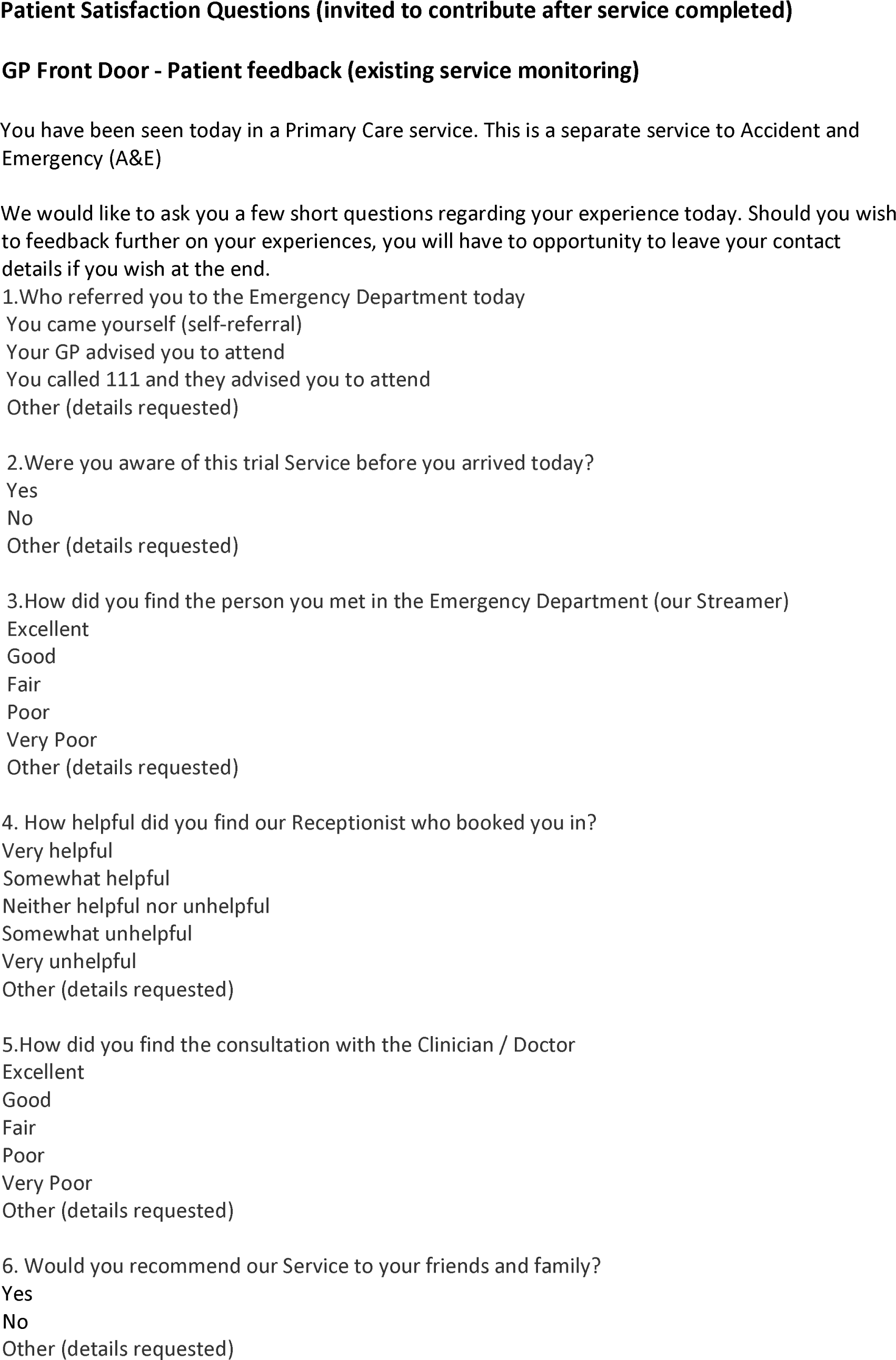
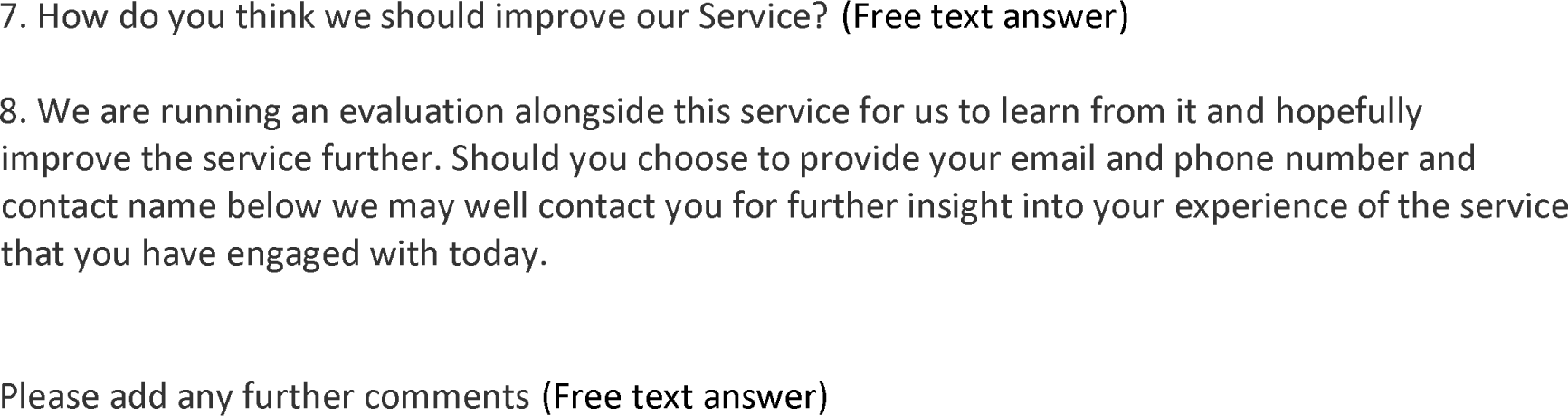

## Notes

### Competing Interest Statement

The authors have declared no competing interest.

